# Early effects of a novel 5-HT_4_ agonist (PF-04995274) and the SSRI citalopram on emotional cognition in unmedicated depression: the RESTAND study

**DOI:** 10.1101/2025.08.28.25334462

**Authors:** A. Gillespie, A.N. de Cates, J. Scaife, M. Blandhol, M.A.G. Martens, D Gibson, B.R Godlewska, W Howard, P.J. Cowen, S.E. Murphy, C.J. Harmer

**Affiliations:** University Department of Psychiatry, Warneford Hospital, University of Oxford, Oxford, UK; Oxford Health NHS Foundation Trust, Warneford Hospital, Oxford, UK; Institute for Mental Health, University of Birmingham, Edgbaston, Birmingham, UK

## Abstract

**Background:** Selective serotonin reuptake inhibitors (SSRIs) are limited by inadequate response in a significant minority of patients, slow onset, minimal cognitive benefit, and side effects. Preclinical studies suggest selective serotonin 4 receptor (5-HT_4_R) agonists may produce faster antidepressant effects via distinct mechanisms, however there has been no experimental research in clinical populations to date.

**Aims:** To test whether the novel 5-HT_4_R partial agonist PF-04995274 produces early behavioural and neural changes in emotional cognition similar to SSRIs in patients with unmedicated major depressive disorder (MDD).

**Method:** In a double-blind, placebo-controlled trial, 90 participants with MDD were randomised to 7 days of PF-04995274 (15 mg), citalopram (20 mg), or placebo. Emotional processing was assessed using a behavioural facial expression recognition task and fMRI of implicit emotional face processing (days 6–9). Observer– and self-reported symptoms of depression were also measured at baseline and study end.

**Results:** As anticipated, citalopram reduced accuracy and reaction time for negative faces, with corresponding fMRI changes (reduced left amygdala activation to emotional faces and valence-specific shifts in cortical regions). In contrast, PF-04995274 produced no change in behavioural negative bias or amygdala activity but increased medial-frontal cortex activation across valences. While this was not a clinical trial, both active treatments reduced observer-rated depression severity relative to placebo; PF-04995274 also reduced self-reported depression, state anxiety, and negative affect. No major adverse events occurred.

**Conclusions:** PF-04995274 was not associated with the typical antidepressant profile of negative bias reductions seen with citalopram but was associated with distinct medial-frontal activation during an emotional faces task and displayed preliminary evidence of early clinical improvement, suggesting a potential alternative mechanism for antidepressant effects. Findings support further clinical trials of 5-HT4R agonists and investigation of pro-cognitive and mood effects.

**Clinicaltrials.gov registration number:** NCT03516604.

**Data set information:** Analysis scripts and selected data will be available on publication.

## Introduction

Selective serotonin reuptake inhibitors (SSRIs) are the most commonly prescribed antidepressants for major depressive disorder (MDD). First line treatment with SSRIs effectively reduces depressive symptoms in approximately half of patients with depression, but many others experience an insufficient response(1). SSRIs are also limited by a slow onset of therapeutic effect(2), common side effects such as sexual dysfunction(3), and limited benefit for cognitive impairment(4). Considering the global prevalence of MDD(5), identifying alternative treatments to tackle these limitations is essential. A key mechanism of SSRI action is their ability to ameliorate negative biases in information processing common to MDD(6), such as greater recall of negative (vs positive) information and a greater likelihood of interpreting ambiguous stimuli negatively(7). The SSRI citalopram, for example, reduces negative biases in the recall of self-referential words and the interpretation of facial expressions(8), and attenuates neural response to negative faces in the amygdala and associated networks(9). These changes occur early in treatment, are associated with later symptom improvement, and are seen across several existing antidepressant treatments with different pharmacology, suggesting they may reflect a common or converging mechanism (10,11). Emotional processing models have therefore been used in antidepressant drug development as an early biomarker of antidepressant action(12).

One approach for developing novel antidepressant treatments is selective targeting of serotonin receptor subtypes involved in mood and cognition. Of particular interest is the potential of selective serotonin 4 receptor (5-HT_4_R) agonists in the treatment of depression and cognition(13). Positron emission tomography (PET) studies have shown reduced 5-HT_4_R binding in MDD(14) and preclinical work demonstrates that 5-HT_4_R agonists produce antidepressant– and anxiolytic-like effects in rodent models, including the forced-swim test, sucrose preference, and elevated plus maze (15–17). Notably, these effects consistently occur more rapidly with 5-HT_4_R agonism than with SSRIs, which may be due to a rapid induction of neuroplastic changes (2-3 times faster than with SSRIs) and the direct excitation of serotonin cells in the dorsal raphe nucleus. Together, these findings suggests that the 5-HT_4_R is a promising fast-acting antidepressant target.

### Aim of current study

The current experimental medicine study aimed to investigate the translation of these preclinical antidepressant effects of 5-HT_4_R agonism to human models of antidepressant action, in participants with un-medicated major depressive disorder. Specifically, we used a novel, highly selective 5-HT_4_R partial agonist (PF-04995274) to investigate early effects (6-9 days) on behavioural and neuroimaging models of emotional cognition previously shown to be sensitive to the effects of SSRIs. We hypothesised that PF-04995274 would reduce negative emotional biases and reduce the neural response to negative stimuli, similar to the effects seen with SSRIs. We included a group randomised to citalopram as a positive control to confirm expected SSRI-induced changes. As this was the first investigation of 5-HT_4_R agonism in MDD, we also measured effects on depression symptomatology.

## Methods

### Participants

Between August 2018 and July 2022, ninety participants (aged 18-61) with Major Depressive Disorder (DSM-5) were recruited. Diagnosis was confirmed by study psychiatrists, using the Structured Clinical Interview for DSM Disorders (SCID). Participants had received no drug or face-to-face psychological treatment for their depression within the previous six weeks, and were not taking any other psychotropic medication. Participants were adults of any gender, fluent in English, with no contraindications to 5-HT_4_R agonism or citalopram. We excluded potential participants who had failed to respond to antidepressant medication in their current episode of depression, or were at clinically significant risk of suicide. We also excluded any potential participant who may be at increased risk of adverse events (e.g., severe gastro-intestinal problems). See Supplementary Materials for a full list of inclusion and exclusion criteria.

The authors assert that all procedures contributing to this work comply with the ethical standards of the relevant national and institutional committees on human experimentation and with the Helsinki Declaration of 1975, as revised in 2013. All procedures involving human subjects/patients were approved by the South-Central NHS Research Ethics Committee (18/SC/0076). The protocol was pre-registered with clinicaltrials.gov (NCT03516604). Participants gave written and verbal informed consent. Participants were paid £200 for their participation (or £170 if the MRI scan was omitted due to contraindication).

### Design

This was a between-subject, double-blind, randomised, placebo-controlled experiment. Participants received 7 days of PF-04995274 (15 mg once daily), citalopram (20 mg once daily), or placebo, taken orally; up to two additional days’ dosing was allowed to accommodate scheduling.

The study included four visits in total: (a) Screening – Interview and Physical Health Screen; (b) Dosing Visit; (c) Research Visit One (including fMRI scan) on day 6-9, and (d) Research Visit Two (including all cognitive tasks and end of study symptom assessment) on day 7-9. By day 6 when scanning took place, steady state plasma levels were expected; the half-life (t1/2) of PF-04995274 is ∼30 hours, and citalopram is ∼35 hours. All visits occurred at the Warneford Hospital, Oxford University Department of Psychiatry. Scanning took place at the Oxford Centre for Human Brain Activity (OHBA), part of the Oxford Centre for Integrative Neuroimaging (OxIN) (previously Wellcome Centre for Integrative Neuroimaging, WIN).

### Interventions

PF-04995274 is a highly-selective 5-HT_4_R partial agonist, developed by Pfizer, and provided (alongside matched placebo tablets) via the MRC-Industry Asset Sharing Initiative. It has been previously tested in human clinical trials of healthy volunteers, with PET studies establishing that 5mg of PF-04995274 produced > 80% receptor occupancy of brain 5-HT_4_R four hours after the first dose, and that 15mg was well-tolerated (Pfizer, 2011a).

Citalopram is a selective-serotonin reuptake inhibitor (SSRI), and is a safe, well-tolerated, licensed treatment for MDD. It was provided and encapsulated (alongside matched placebo capsules) by Cardiff and Vale University Health Board, St. Mary’s Pharmaceutical Unit (SMPU).

The first dose was administered on site, with participants monitored for 3 hours post-dosing. All subsequent administration occurred at home.

### Randomisation and blinding

The research team and participants were blinded to group allocation. The randomisation code (stratified by gender) was drawn up and held by Oxford Health Foundation Trust Pharmacy (Clinical Pharmacy Support Unit – CPSU – Kennington), using an online randomisation tool (Sealed Envelope); they also stored and dispensed the medication. Group allocation was concealed using sequential numbered containers (see Supplementary Materials for more details).

### Power calculation

Based on previous citalopram–placebo comparisons (18), 19 participants per group would provide 90% power (α = 0.05) for behavioural measures. To accommodate potential exclusions, subtler effects in fMRI analyses, and the less well-characterised effects of 5-HT4R agonism, we aimed for 25 participants with complete behavioural and fMRI data per group. Including 13 participants who completed behavioural assessments but not MRI (due to contraindications), 90 participants were recruited.

### Clinical assessment and questionnaire measures

To obtain an observer-related measure of depression severity, trained researchers assessed participants at baseline and end-of-study using the 17-item Hamilton Rating Scale for Depression (HAM-D)(19).

To obtain a self-report measure of depression severity, participants completed the Beck Depression Inventory-II(20) at baseline and end-of study, as well as the Snaith-Hamilton Pleasure Scale (SHAPS)(21) to measure anhedonic symptoms. At baseline, the Spielberger State-Trait Anxiety Inventory, Trait Version (STAI-T)(22) and Eysenck Personality Questionnaire(23) were also completed, to assess baseline trait anxiety and personality. State affect and anxiety were measured at both research visits using: the Positive and Negative Affect Scale (PANAS)(24); visual analogue scales (VAS)(25); and the Spielberger State-Trait Anxiety Inventory, State Version (STAI-S)(22). Commonly reported side effects were also measured before dosing, at each visit, and at home across the study period. At the end of the study, participants were asked to guess their group allocation with a multiple-choice question.

### Behavioural measure of emotional cognition

Emotional cognition was assessed at Research Visit Two (day 7-9) with a battery of cognitive tasks from the Emotional Test Battery (P1vital® Limited Products)(26), including the Facial Expression Recognition Task (FERT): Participants viewed briefly presented (500 ms) faces depicting basic emotions (happiness, fear, anger, disgust, sadness, surprise, neutral) at varying intensities and were asked to classify each via button press. There were 250 trials in a fixed pseudorandomized order. (See Supplementary Materials for details of other emotional cognition tasks. Results from non-emotional cognition tasks are reported elsewhere (27)).

### Neuroimaging measure of emotional cognition

Neural activity during emotional processing was assessed using an implicit emotional cognition task conducted during an fMRI scan. Briefly, emotionally-valenced faces (fearful or happy) were presented on screen for 100ms. Participants were instructed to indicate the assumed gender of the face (male or female) as quickly as possible using a button press. No reference was made to face emotion during the instructions. There was a rest block at the start of the experimental run, and then the task followed an A-B-Rest design, where a block of condition A (fearful faces, 18s) was followed by a block of condition B (happy faces, 18s) then a block of rest (12s). Seven repetitions yielded 126s per emotional condition and 96s rest. This version was a recent modification of the task, which we have shown to be sensitive to the acute effects of antidepressants on neural processing(28). The task software was written using Psychopy version 1.84.2.

Blood-oxygenation-level-dependent (BOLD) fMRI and T1-weighted anatomical images were acquired using a 3-Tesla Siemens Prisma scanner, equipped with a 32-channel head matrix coil (Siemens, Erlangen, Germany). Foam padding and a head restraint were used to control head movement. In the scanner, participants also completed a memory encoding task and resting-state scan (reported elsewhere (27)), and an arterial spin-labelling (ASL) scan. The full acquisition protocol, along with radiography procedure, is available from the Open WIN MR Protocols database here: <u>10.5281/zenodo.6107724</u>.

## Statistical analysis

### Behavioural and clinical data

For all behavioural data, outliers were determined based on blinded visual inspection of histogram plots. Outlier removal/sensitivity checks were determined on 12^th^ January 2023, prior to unblinding. There was determined to be one outlier participant in the FERT data, who was removed during sensitivity checks.

Demographic characteristics and baseline clinical measures are reported descriptively. For all analyses, the citalopram group was first compared to the placebo group to confirm expected SSRI effects. The 5-HT_4_R group was then compared to placebo to determine the effect of PF-04995274. Group differences in cognitive task performance and clinical outcomes were assessed using ANOVA, with group as between-participant factor and task condition (e.g. valence) as within-participant factor. For self-report scales where baseline measures were collected (HAM-D, BDI and SHAPS), baseline severity was included as a covariate. Chi-square tests were used to assess group differences in number of reported side-effects. A p value less than 0.05 was used to denote statistical significance. Partial eta-squared is reported as a measure of effect size. R (version 4.3.3) was used for all data processing and analysis.

As the STAI-S, PANAS and VAS measures during research visits were not completed an equal number of times for all participants (due to differences in whether participants were suitable for MR scanning and completed one or two research visits), an average score was calculated for each participant to indicate overall end-of-study affect.

### Magnetic Resonance Imaging Data

fMRI data were pre-processed and analysed using FEAT (FMRI Expert Analysis Tool), version 6.0.4, part of FSL (FMRIB’s Software Library2). See Supplementary Material for pre-processing steps.

In the first-level analysis, individual activation maps were computed using the general linear model with local autocorrelation correction. Three explanatory variables were modelled: “happy” and “fear” images and explicit fixation cross, to replicate the models in our previous papers(28). Temporal derivatives were included in the model. Variables were modelled by convolving each block with a haemodynamic response function with a standard deviation of 3s and a mean lag of 6s. Two participants had significant movement (one allocated to citalopram, one allocated to placebo) and were therefore excluded from analysis. All other absolute displacements were less than 2.2 voxels and relative displacements less than 0.2 voxels. FSL motion outliers tool was used to reduce the influence of remaining motion. At the whole-brain level, we contrasted fearful images with happy and fixation cross: (1) fear > fixation; (2) happy > fixation; (3) fear > happy; (4) happy > fear; (5) mean (of happy and fearful faces) > fixation.

In the second-level analysis, whole-brain individual data were combined at a group level (placebo vs. active drug – either citalopram or PF-04995724, our 5-HT_4_R agonist) using a mixed-effects group analysis across the whole brain corrected for multiple comparisons. Groups were contrasted with each other using the following comparisons: (1) placebo > active; (2) active > placebo; (3) mean of active + placebo participants. Brain activations showing significant group differences were identified using cluster-based thresholding (Z > 3.1, p < 0.05 FWE corrected). Significant interactions from whole-brain analyses were further explored by extracting percentage BOLD signal change with Featquery for each type of contrast.

As the amygdala(29) and prefrontal cortex(30) were a particular focus of our hypothesis, the amygdala, medial frontal cortex (MFC), and orbitofrontal cortex (OFC) were pre-specified as regions of interest (ROI). A functional ROI mask was created for each by multiplying mean activation (mean>fixation contrast) for all participants (placebo, citalopram and PF-04995274) by the Harvard-Oxford subcortical atlas anatomical mask at a 50% threshold. Percentage BOLD signal change for each contrast in each hemisphere was then extracted to identify the profile of drug effect.

## Results

### Sample demographics

90 participants completed the full period of study medication and the final research visit. 77 of these had no scanning contraindications and completed an MRI scan visit prior to their final research visit. Participants were predominantly white, native English speakers aged 20-40, with education equivalent to completion of an undergraduate degree (Table 1).

**Table 1:** Demographics and baseline self-report – All participants.

| Characteristic | 5HT<sub>4</sub>, N = 31<sup>1</sup> | Citalopram, N = 30<sup>1</sup> | Placebo, N = 29<sup>1</sup> |
| --- | --- | --- | --- |
| Sex |  |  |  |
| F | 19 (61%) | 19 (63%) | 17 (59%) |
| M | 12 (39%) | 11 (37%) | 12 (41%) |
| Age (decimal) | 36.70 (13.10) | 34.81 (12.06) | 32.48 (10.33) |
| Ethnicity (grouped) |  |  |  |
| White | 27 (87%) | 22 (73%) | 24 (83%) |
| Other | 4 (13%) | 8 (27%) | 5 (17%) |
| First language - English |  |  |  |
| Y | 19 (61%) | 20 (67%) | 23 (79%) |
| N | 12 (39%) | 10 (33%) | 6 (21%) |
| Years of Education | 16.71 (2.69) | 16.90 (3.51) | 17.30 (2.47) |
| BMI | 24.97 (4.41) | 25.15 (3.69) | 25.51 (4.16) |
| Handedness - Edinburgh Inventory | 18.76 (2.24) | 17.43 (3.76) | 18.03 (3.08) |
| Extraversion (EPQ) | 8.52 (4.77) | 8.93 (4.49) | 9.07 (5.36) |
| Neuroticism (EPQ) | 17.52 (4.25) | 16.33 (4.18) | 17.76 (4.14) |
| Psychoticism (EPQ) | 4.45 (2.90) | 4.00 (2.73) | 3.38 (2.60) |
| Social desirability (EPQ) | 7.58 (3.50) | 7.30 (3.31) | 8.31 (4.83) |
| Trait anxiety (STAIT) | 58.65 (9.19) | 57.77 (7.75) | 59.31 (7.26) |
| Self report depressive symptoms (BDI) | 24.16 (10.83) | 24.45 (9.43) | 24.86 (10.50) |
| Anhedonic symptoms (SHAPS) | 4.32 (2.76) | 3.83 (3.00) | 3.83 (3.06) |
| Hormonal contraception (% of female participants) |  |  |  |
| No hormonal contraception | 13 (68%) | 12 (63%) | 9 (53%) |
| Oral contraceptive pill - hormonal | 4 (21%) | 1 (5.3%) | 6 (35%) |
| Intra uterine system (IUS) - hormonal | 1 (5.3%) | 5 (26%) | 0 (0%) |
| Unknown | 1 (5.3%) | 1 (5.3%) | 2 (12%) |

<sup>1</sup>n (%); Mean (SD)
*Ethnicity, other, self-defined: 5HT<sub>4</sub> = Chinese, Eastern European, Askanazi Jewish; Citalopram = Asian (n=4), Black, Mixed, Mixed Indian and British; Placebo = Asian, Thai, Chinese, Mixed, Mixed Welsh/Indo guaynese*

### Blinding

There was a significant association between medication group and guessed medication (*X^2^* (6, N=90)=12.92, p=0.04), indicating some level of un-blinding; this was driven by the citalopram group (citalopram vs placebo, *X^2^* (3, N=59)=8.09, p=0.04; PF-04995274 vs placebo, *X^2^* (3, N=60)=2.06, p=0.56). 43.34% of participants allocated to citalopram correctly guessed their allocation (vs 9.78% of those allocated to PF-04995274, and 37.93% of those allocated to placebo), and 60% correctly guessed they had an active drugs (vs 25.81% of PF-04995274 participants).

### Changes in observer-rated symptoms

At baseline, the mean HAM-D score for the whole sample was 16.1 (sd=2.78), indicating mild to mod erate depression(30). Groups were reasonably well matched, with marginally lower severity in the ci talopram group (m=15.2, sd=2.8) compared with the placebo (m=16.9, sd=2.88) and PF-04995274 (m =16.2, sd=2.51) groups.

Controlling for baseline severity, the citalopram group scored significantly lower on the HAM-D at the final research visit (m=12.7, sd=5.20) compared to placebo (m=15.5, sd=3.9) (F(1, 54)=5.50, p=0.02, ηp2 = 0.09), as did the PF-04995274 group (m=12.7, sd=4.04) (F(1, 56)=7.61, p=0.008, ηp2 = 0.12) (**Figure 1**Figure 3). The significance and size of these effects remained, or increased in sensitivity analyses (removing sleep-related items, using core six items of the HAM-D only^1^, removing potential outlier participants).

**Figure 1.**
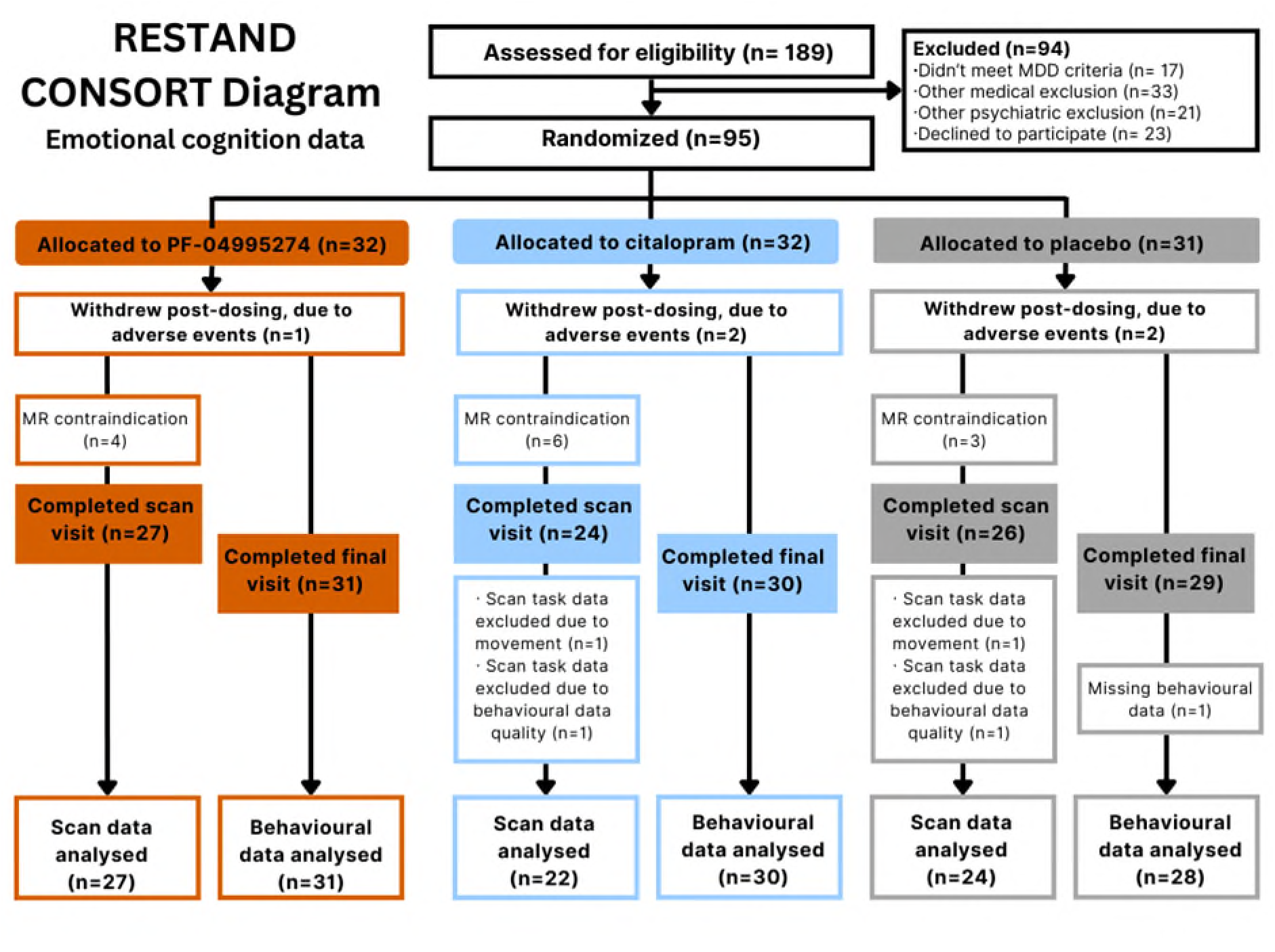
RESTAND CONSORT Diagram. MDD = Major Depressive Disorder. MR = Magnetic Resonance. Grey = placebo, Blue = citalopram, Orange = PF-04995274.

**Figure 2.**
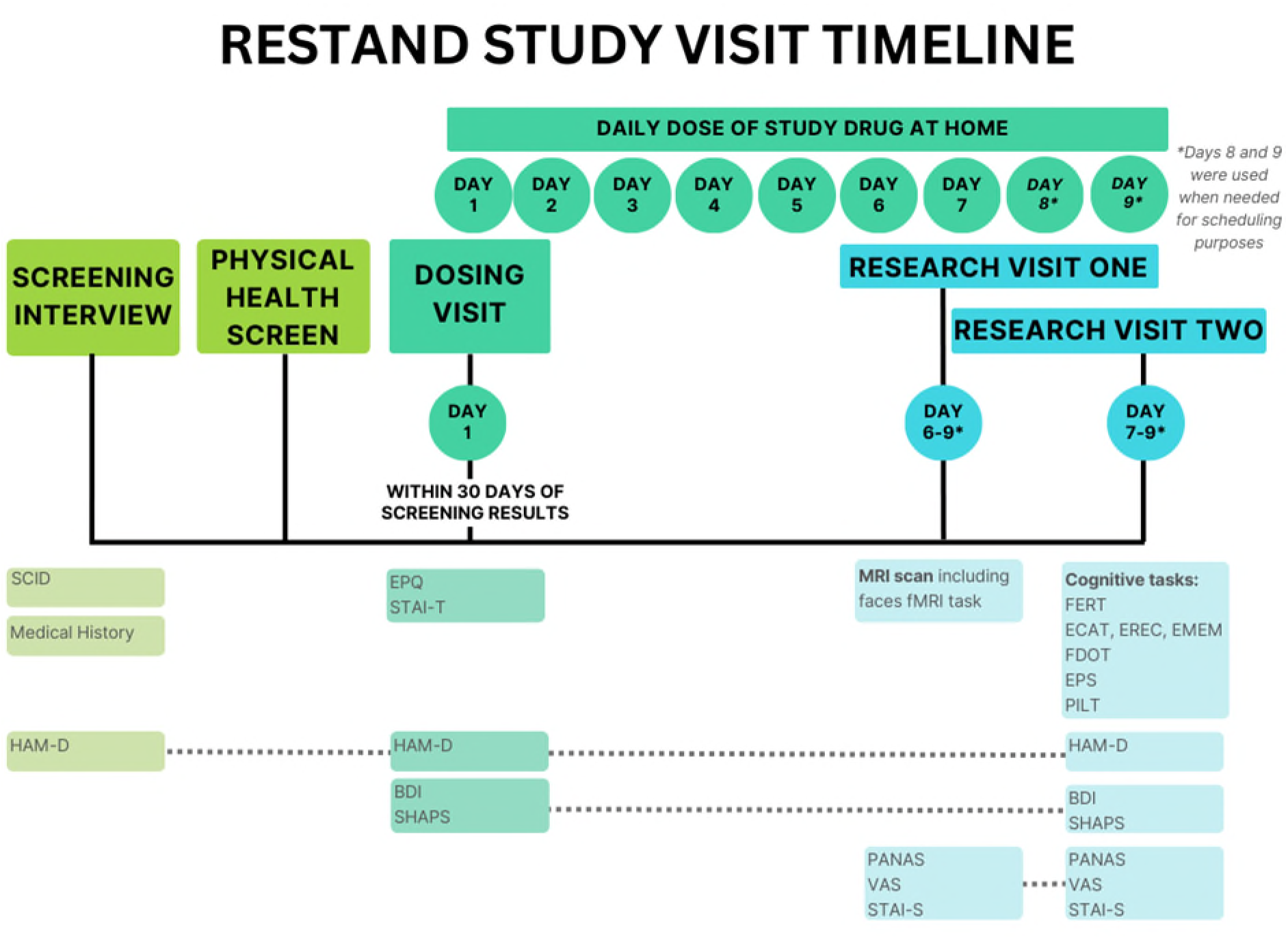
RESTAND Study Visit Timeline. BDI = Beck Depression Inventory. ECAT = Emotional Categorisation Task. EMEM = Emotional Recognition Memory Task. EPQ = Eysenck Personality Questionnaire. EPS = Emotion Potentiated Startle task. EREC = Emotional Recall Task. FDOT = Faces Dot Probe Task. FERT = Facial Expression Recognition Task. HAM-D = Hamilton Depression Rating Scale, an observer-rated measure of depression severity. MRI = Magnetic Resonance Imaging. PANAS = Positive and Negative Affect Schedule. PILT = Probabilistic Learning Task. SCID = Structured Clinical Interview for DSM-5. SHAPS = Snaith-Hamilton Pleasure Scale. STAI-S = State Trait Anxiety Inventory, State version STAI-T = State Trait Anxiety Inventory, Trait version. VAS = Visual Analogue Scales.

**Figure 3.**
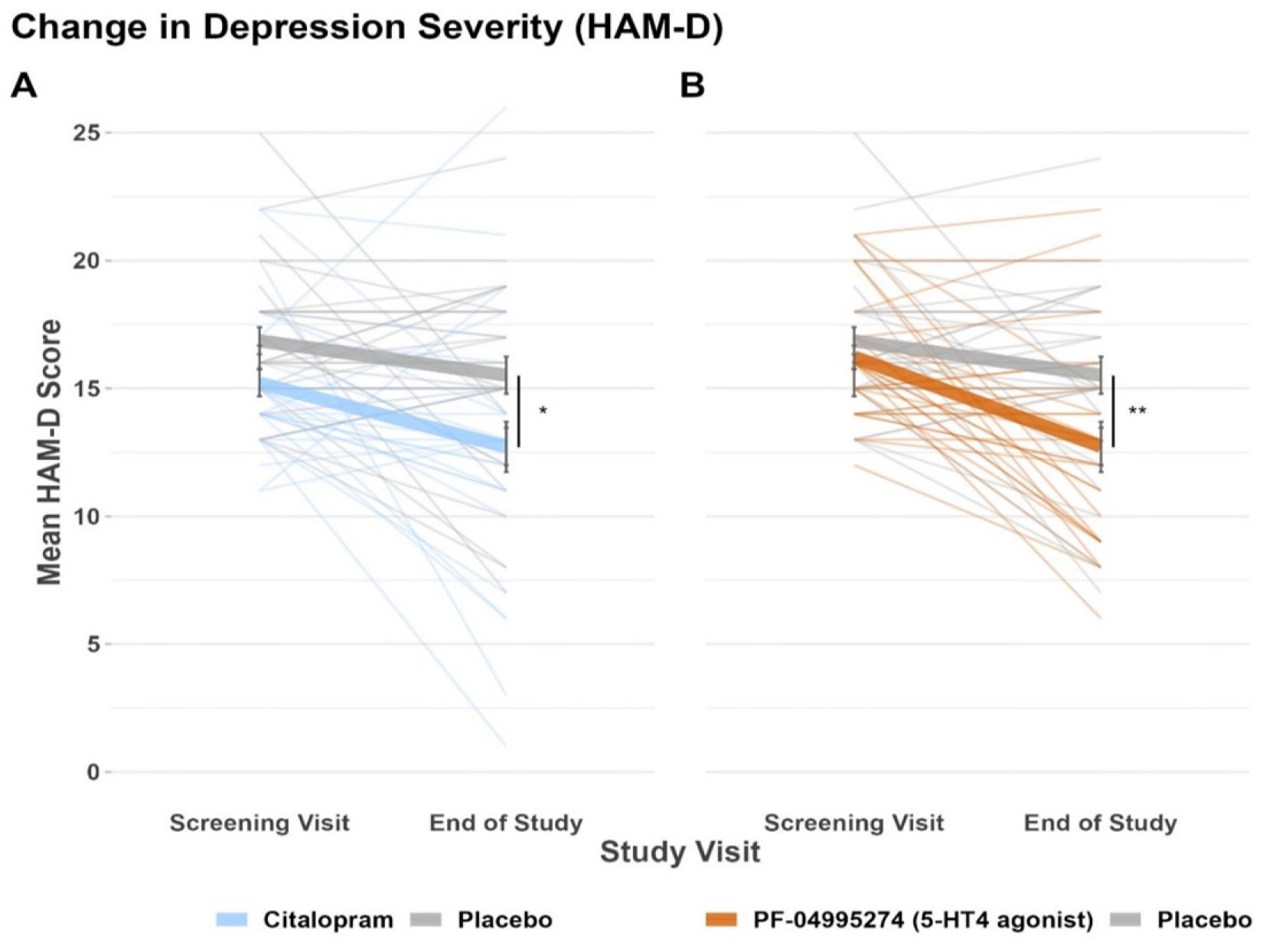
Change in Depression Severity (HAM-D). HAM-D = Hamilton Depression Rating. **A** – citalopram group vs placebo group. **B** – PF-04995274 group vs placebo group. * = p < 0.05, ** = p< 0.01 for main effect of group on ANOVA, controlling for baseline score. Errors bars show standard error of the mean. Grey = placebo, Blue = citalopram, Orange = PF-04995274.

### Changes in self-reported symptoms and affect

Controlling for baseline severity, the PF-04995274 group – but not the citalopram group – reported significantly lower depressive symptoms (BDI) at the final visit than placebo (**Table 2** for all statistics). Similarly, only the PF-04995274 group reported lower state anxiety (STAI-S) than placebo at the final visit. Both groups reported significantly lower negative affect at final visits compared to placebo, with double the effect size in the PF-04995274 vs citalopram group. Only the citalopram group reported significantly lower positive affect at final visits compared to placebo. Neither group significantly differed from placebo in reported levels of anhedonia (SHAPS) – when controlling for baseline severity – or on any visual analogue scales (see Supplementary Materials).

**Table 2:** End of study self-report – All participants.

| Characteristic | PF-04995274, N = 31<sup>1</sup> | Citalopram, N = 30<sup>1</sup> | Placebo, N = 29<sup>1</sup> | F statistic | P value | Partial eta squared (η<sup>2</sup>) |
| --- | --- | --- | --- | --- | --- | --- |
| Depressive symptoms (BDI)<sup>+</sup> | **17.69 (11.38)** | 20.63 (10.82) | **22.38 (9.28)** | Citalopram vs. placebo - F(1,52)=0.44<br>**PF-04995274 vs. placebo - F(1,55)=6.33** | 0.5<br>**0.015** | 0.008<br>0.1 |
| Anhedonic symptoms (SHAPS)<sup>+</sup> | 3.31 (2.71) | 3.93 (2.42) | 3.45 (2.81) | Citalopram vs. placebo – F(1,52)=0.87<br>PF-04995274 vs. placebo - F(1,54)=0.05 | 0.36<br>0.82 | 0.02<br>0.001 |
| State anxiety across final research visit (STAI-S) | **42.46 (9.70)** | 44.17 (10.82) | **45.01 (9.77)** | Citalopram vs. placebo - F(1,267)=0.31<br>**PF-04995274 vs. placebo - F(1,273)= 3.92** | 0.58<br>**0.049** | 0.001<br>0.02 |
| Positive affect across final research visit (PANAS - P) | 21.47 (7.35) | **20.18 (5.14)** | **22.03 (6.63)** | **Citalopram vs. placebo – F(1,267)=7.29**<br>PF-04995274 vs. placebo - F(1,273)=0.42 | **0.007**<br>0.52 | 0.02<br>0.002 |
| Negative affect across final research visit (PANAS - N) | **14.80 (4.80)** | **15.54 (5.60)** | **16.99 (5.82)** | **Citalopram vs. placebo - F(1,267)=4.93**<br>**PF-04995274 vs. placebo - F(1,273)=10.75** | **0.027**<br>**0.001** | 0.02<br>0.04 |

<sup>1</sup>Mean (SD). BDI = Beck's Depression Inventory. PANAS = Positive and Negative Affect Schedule. P = Positive subscale. N = Negative subscale. SHAPS = Snaith-Hamilton Pleasure Scale. STAI-S = State Trait Anxiety Inventory. <sup>+</sup>HAM-D and BDI analyses controlled for baseline severity.

### Side effects

PF-04995274 group reported new fatigue significantly more often (X^2^=6.5, p=0.04), all reports of which were described as mild-moderate. There were no other significant group differences in new reported side effects (p>0.1, see Supplementary Materials).

Three participants (one on placebo, one on citalopram, one on PF-04995274) withdrew from the study due to adverse events, all of which resolved within 24 hours of discontinuation.

### Behavioural measure of emotional cognition – Facial Expression Recognition Task^2^

When comparing citalopram to placebo, there was no main effect of group (F(1,56)=0.49, p=0.49, ηp2=0.009) but there was a significant interaction between medication group and valence on total percentage accuracy of emotional face classification (F(1,56)=4.30, p=0.043, ηp2 =0.07). Consistent with previous reports, the citalopram group showed greater accuracy for positive faces compared to placebo, and reduced accuracy for negative faces (**Figure 4** A). When exploring the misclassification rates i.e. whether medication group was associated with differences in misclassification of faces as positive, neutral, or negative emotions, there were no significant associations but a sensitivity analysis (potential outlier removal) indicated a trend for the citalopram group having a reduced negative misclassification rate compared to placebo (F(1,55)=3.056, p=0.086). There was no significant difference between citalopram and placebo group in positive or neutral misclassification rate (p>0.12).

**Figure 4.**
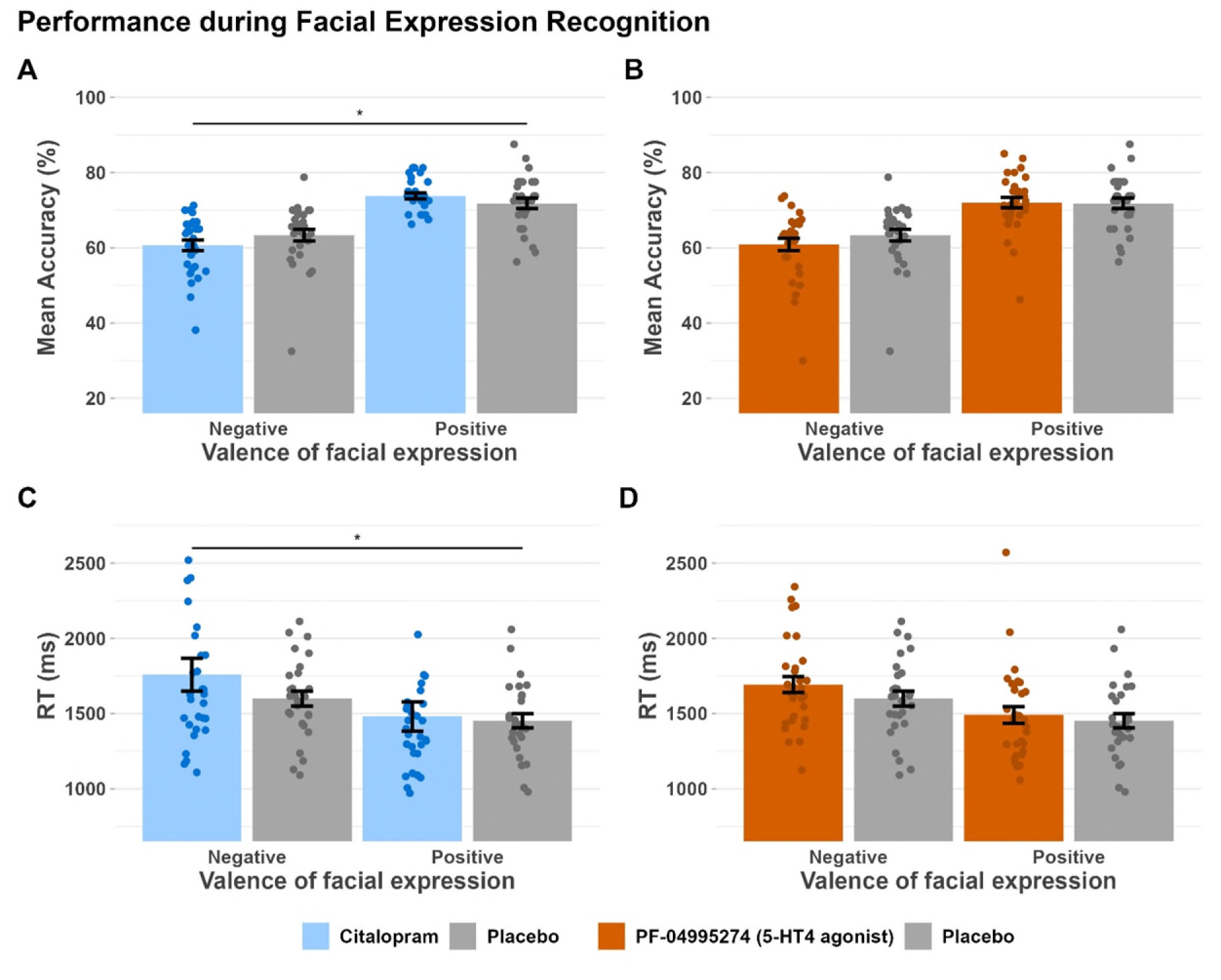
Performance during Facial Expression Recognition Task. **A** – citalopram group vs placebo group, comparing percentage accuracy for negative vs positively valenced faces. **B** – PF-04995274 group vs placebo group, comparing percentage accuracy for negative vs positively valenced faces. **C** – citalopram group vs placebo group, comparing reaction time for negative vs positively valenced faces. **D** – PF-04995274 group vs placebo group, comparing reaction time for negative vs positively valenced faces * = p < 0.05, for interaction between group and valence on ANOVA. Errors bars show standard error of the mean. Grey = placebo, Blue = citalopram, Orange = PF-04995274.

Similarly, when comparing reaction times for the citalopram group to placebo, there was no main effect of group (F(1,56)=0.98, p=0.325, ηp2=0.02) but there was a significant interaction between group and valence on speed of emotional face classification (F(1,56)=6.78, p=0.012, ηp2=0.11), with the citalopram group showing slower reaction times for negative faces (**Figure 4** C).

When comparing the PF-04995274 group to placebo, there was no main effect of group on accuracy (F(1,57)=0.76, p=0.39, ηp2=0.01) or reaction time (F(1,57)=1.14, p=0.289, ηp2=0.02), and no interaction between group and valence for accuracy (F(1,57)=1.09, p=0.301, ηp2=0.02) or reaction time (F(1,57)=1.39, p=0.244, ηp2=0.02) (**Figure 4** B, D). There was no significant difference in negative, neutral or positive misclassification rate (p>0.4).

The significance, size and pattern of all results remained consistent in all sensitivity checks (using an adjusted accuracy measure which accounts for response bias, transforming the data to address skew, removing a potential outlier participant).

### Neuroimaging measure of emotional cognition – fMRI Emotional faces task

For behavioural data (accuracy and reaction time for gender discrimination) and broad task effects in the fMRI data, see Supplementary Materials.

### Whole brain – medication effects

When comparing citalopram to placebo, there was no difference for mean of emotional faces (fear+happy) vs fixation, but there was significant differential activation for fear vs happy in six clusters covering angular gyrus, lateral occipital cortex, cerebellum, middle temporal gyrus and fusiform gyrus. In all clusters, the placebo group showed greater activation for fearful faces and the citalopram group showed greater activation for happy faces (p<0.01, see **Figure 5** and Supplemental Materials). When comparing the PF-04995274 group to placebo, there were no significant differences on any contrast.

**Figure 5.**
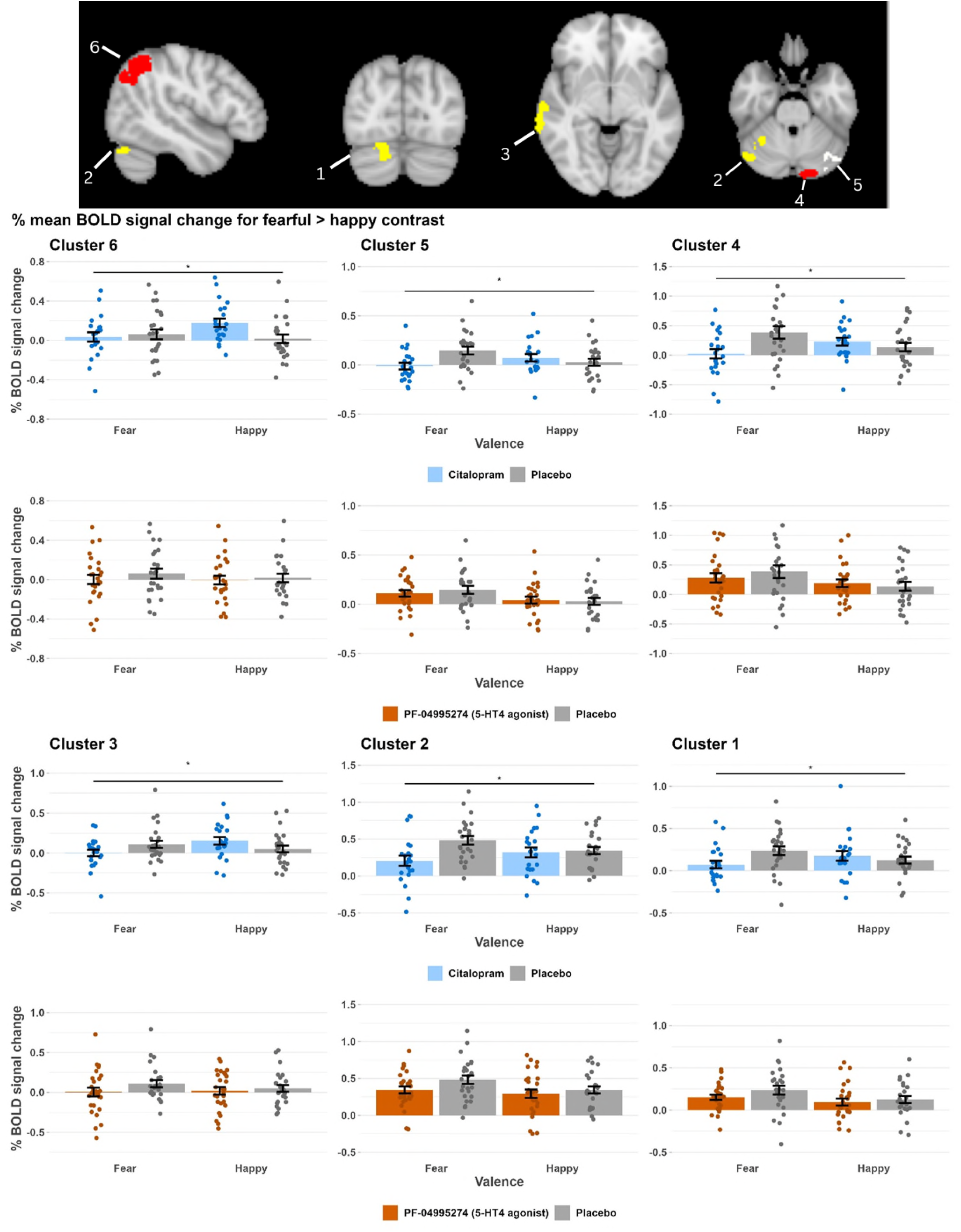
% mean BOLD signal change in 6 clusters with a significant difference between citalopram and placebo group for fearful > happy contrast. Cluster 1 = occipital fusiform gyrus, lingual gyrus. Cluster 2 = cerebellum, occipital fusiform gyrus, lateral occipital. Cluster 3 = middle temporal gyrus (posterior), superior temporal gyrus. Cluster 4 = cerebellum, occipital fusiform, cerebral cortex. Cluster 5 = cerebellum. Cluster 6 = angular gyrus and lateral occipital. * = p < 0.05, ** = p < 0.01 for interaction between group and valence on ANOVA. Errors bars show standard error of the mean. Grey = placebo, Blue = citalopram, Orange = PF-04995274.

### ROI analyses

In the left amygdala, when comparing citalopram to placebo, there was a main effect of group (F(1,90)=4.008, p=0.048, ηp2=0.04), but no significant interaction between group and face valence on % BOLD signal change (F(1,90)=0.001 p=0.97, ηp2=0.0001) i.e. the citalopram group showed reduced amygdala BOLD response across valences (**Figure 6**). There was no main effect or interaction for the right amygdala (p>0.4). When comparing the PF-04995274 group to placebo, there was no significant main effect or interaction (p>0.15) for the left amygdala, nor the right amygdala (p>0.08).

**Figure 6:**
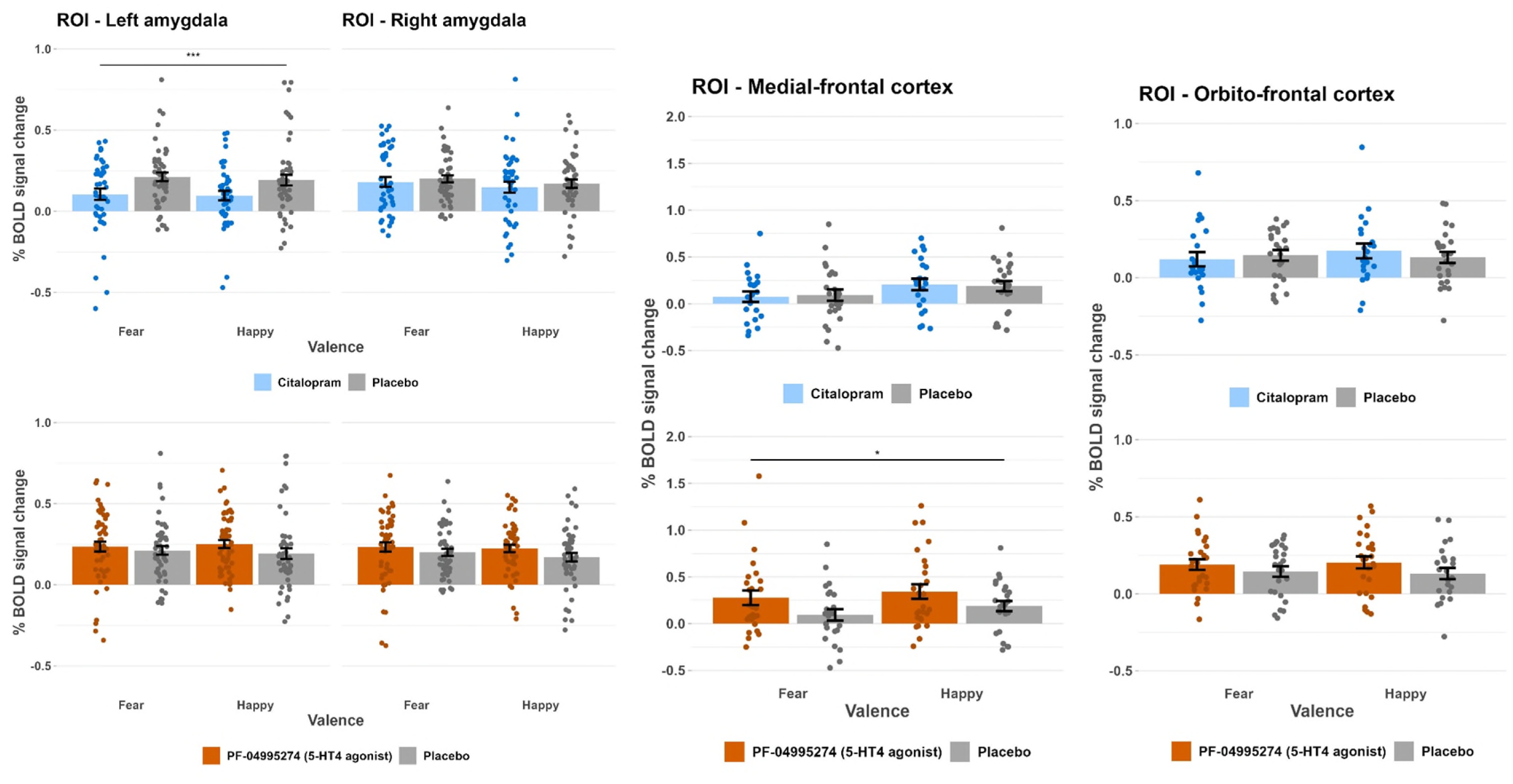
% mean BOLD signal change in left and right amygdala, medial frontal cortex, and orbito-frontal cortex, defined using functional mask. Values extracted separately for fearful > fixation and happy > fixation, comparing citalopram group vs placebo group and then PF-04995274 group vs placebo group. *** = p < 0.001, for interaction between group and valence on ANOVA. Errors bars show standard error of the mean. Grey = placebo, Blue = citalopram, Orange = PF-04995274.

In the orbito-frontal cortex, there was no significant difference for any group comparison, on any contrast (**Figure 6**).

In the medial-frontal cortex, there was a main effect of group when comparing the PF-04995274 group to placebo (F(1,100)=5.98, p=0.016, ηp2=0.06), but no significant interaction between group and face valence (F(1,100)=0.042, p=0.84, ηp2=0.0004) i.e. the PF-04995274 group showed greater BOLD response across valences (**Figure 6 Error! Reference source not found.**). There was no main effect or interaction when comparing citalopram group to placebo (p>0.7).

## Discussion

The primary result of this double-blind, randomised study is that unmedicated depressed participants given a novel 5-HT_4_R agonist (PF-04995274) did not display the profile of behavioural and neural changes in emotional cognition seen in participants given citalopram, and anticipated based on previous SSRI research(8,9). However, those given the 5-HT_4_R agonist did show alternative neural changes during emotional cognition and both groups reported significant reductions in depressive symptoms after one week of drug administration compared to placebo.

On a behavioural facial expression recognition task, participants randomised to citalopram showed reduced recognition of – and slower reaction times to – negative faces than the placebo group, with a non-significant trend towards fewer misclassifications of faces as negative, indicative of an amelioration of negative bias. Concurrently, in an implicit emotional faces fMRI task, whole brain analyses revealed significantly reduced activity in response to fearful vs happy faces, compared to placebo, across a number of relevant brain regions including angular gyrus, lateral occipital cortex, cerebellum, middle temporal gyrus, and fusiform gyrus. Additionally, pre-registered ROI analysis showed reduced left amygdala activation to emotionally-valenced faces in the citalopram group. Collectively, these findings align with the general literature on neural effects of SSRIs during emotional processing, which are thought to provide a potential mechanism for SSRI efficacy(6).

In contrast, the 5-HT_4_R agonist group shower no significant differences on any behavioural measures of emotional cognition, valence-specific neural changes, or changes in amygdala BOLD response to emotional stimuli compared to placebo. This broadly aligns with our previous research using 1 or 6 days of prucalopride (1mg), an alternative 5-HT_4_R agonist. However, in ROI analysis, the 5-HT_4_R agonist group did demonstrate significantly increased medial-frontal cortex activation in response to both emotionally-valenced faces. The medial-frontal cortex modulates amygdala activity, and is thought to be involved in higher-level processing of emotional cognition and affective state, including representation of interpersonal relationships (32); interestingly, meta-analytic evidence in studies of patients with MDD/anxiety disorders indicates that psychotherapy such as CBT (but not conventional SSRIs) is typically associated with increased activity in this region(33). This differs from our findings with prucalopride in healthy volunteers, which found 5-HT_4_R agonism was associated with reduced activity in the medial pre-frontal cortex and the inferior parietal lobule during the same task. This may be due to important differences between the compounds; prucalopride has distinct effects on 5-HT_4_R in the gastrointestinal system (28,34), and brain receptor occupancy is less well-established. Alternatively, it may highlight the importance of translating finding into a clinical population; PET studies have already shown that the relationship between 5-HT_4_R function and memory is reversed in healthy volunteers compared to participants with depression(14).

Despite the short duration of the study, participants receiving either active drug scored significantly lower on observer-rated depression severity at the end of the study compared to placebo. In the 5-HT_4_R agonist group, this was accompanied by significant reductions in self-reported depression, state anxiety and negative affect. While the citalopram group had significant unblinding, blinding was well-maintained in the 5-HT_4_R group. Interestingly, the citalopram group reported significant reductions in both negative and positive affect, perhaps suggestive of emotional blunting(35). Our fMRI finding of non-specific reduction in amygdala activation to emotional faces may correspond with this; however, the lack of amygdala differentiation between the valences of the faces across the whole group, as in our previous use of the task (28), may instead indicate that features of this version of the task (namely, more tightly cropped face images) reduce differentiation between fearful and happy expressions.

Interpretating the clinical outcomes should be cautious; this study was not designed as a clinical trial, so measures of depression severity were not our primary outcome and study duration was short. Exclusion criteria were also restrictive and participants had mild-moderate depression severity. Nonetheless, we provide interesting preliminary experimental evidence in a clinical population that 5-HT_4_R agonism displays promise for antidepressant drug development. This is consistent with a recent emulated target trial from our group using large-scale USA electronic health records, which found that prescription of prucalopride is associated with reduced incidence of first ever depression, relative to other anti-constipation agents(36).

Collectively, these results suggest that any antidepressant efficacy of 5-HT_4_R agonism would likely operate via mechanisms distinct from conventional SSRI antidepressants, and should be a focus for future research. In our previous work with prucalopride, we reported pro-cognitive effects of 5-HT_4_R agonism on learning and memory tasks(28,34,37) and our finding of increased medial-frontal activity during emotional cognition may provide additional support for this. One speculation may be that 5-HT_4_R agonism indirectly improves mood by ameliorating cognitive impairment. Pro-cognitive effects, however, remain unexplored in clinical populations, as does any causal link between these and overall clinical improvements.

In conclusion, in the first randomised, experimental study in patients with depression, 6-9 days of 5-HT_4_ R agonism with PF-04995274 produced contrasting changes on behavioural and neuroimaging models of emotional cognition to those seen with conventional SSRI antidepressants, with no indication of reduced negative bias but instead increased medial-frontal activity. Participants allocated to this novel 5-HT_4_ R agonist reported significantly lower depressive symptoms after 7-9 days, a finding which should be confirmed and extended in clinical trials, alongside further experimental work to clarify the distinct underlying mechanisms.

## Data Availability

All behavioural data produced in the present study will be made available online by the end of 2025 and are available upon reasonable request to the authors.

## Acknowledgements

We would like to thank: numerous research assistants for their support with data collection (James Carson, Lucy Wright, Ingrid Martin, Evie Watson) and data processing (Esther Teo and Anutra Guru); numerous medics for support with clinical cover (Michael Browning, Riccardo De Giorgi, Tarek Zghoul, Paul Harrison, Sara Costi); Cassandra Gould Van Praag for her support with the scanning protocol and analysis pipelines; numerous staff (medical, nursing and administrative) at the Oxford Health Clinical Research Facility; the radiographers at the Oxford Centre for Human Brain Activity (OHBA); Orla MacDonald and CPSU (Oxford Health) for their pharmaceutical support; and the Oxford Health BRC Patients Active in Research Group who provided consultation on study adverts and protocol. We would also like to extend a huge thank you to all of the participants who took part in the study, especially as this involved specifically giving their time and energy during ongoing depressive episodes.

## Author Contributions

**ALG** – methodology, investigation, formal analysis, data curation, writing – original draft, writing – review and editing, visualisation, project administration. **ANdeC** – methodology, investigation, data curation, writing – review and editing. **JS** – methodology, investigation, data curation, writing – review and editing. **MB** – investigation, data curation, writing – review and editing. **MAGM** – data curation, software, writing – review and editing. **DG** – investigation, data curation, writing – review and editing. **BG** – methodology, investigation, writing – review and editing. **WH –** writing – review and editing, project administration, resources. **PJC –** funding acquisition, conceptualization, methodology, investigation, writing – review and editing, supervision. **SEM –** funding acquisition, conceptualization, methodology, writing – review and editing, supervision. **CJH** – funding acquisition, conceptualization, methodology, investigation, writing – review and editing, supervision.

## Supplementary Methods

### Complete inclusion criteria

- Male or female;
- Aged 18-65 years;
- Willing and able to give informed consent for participation in the study;
- Sufficiently fluent English to understand and complete the tasks;
- Registered with a GP and consents to GP being informed of participation in study;
- Meet DSM-V criteria for current Major Depressive Disorder [as determined by the Structured Clinical interview for DSM-V (SCID)];
- Participant must have received no drug or face-to-face psychological treatment for the current episode of depression/ in the previous six weeks;
- Participants engaging in sex with a risk of pregnancy must agree to use a highly effective method of contraception from Screening Visit until 30 days after receiving study medication treatment, female participants must not breastfeed, and male participants must not donate sperm. Acceptable methods of contraception include:
  ○ Combined (estrogen and progestogen containing) hormonal contraception associated with inhibition of ovulation; oral, intravaginal or transdermal
  ○ Progestogen-only hormonal contraception associated with inhibition of ovulation: oral, injectable or implantable
  ○ Intrauterine device (IUD)
  ○ Intrauterine hormone-releasing system ( IUS)
  ○ Bilateral tubal occlusion
  ○ Vasectomy (or vasectomised partner)
  ○ Sexual abstinence. Periodic abstinence (calendar, symptothermal, post-ovulation methods), withdrawal (coitus interruptus), and spermicides only are not acceptable methods of contraception).

### Complete exclusion criteria

- History of or current DSM-V bipolar disorder, schizophrenia or eating disorders. Participants who fulfil current criteria for other comorbid disorders may still be entered into the study, if, in the opinion of the Investigator, the psychiatric diagnosis will not compromise safety or affect data quality;
- First-degree relative with a diagnosis of Bipolar Disorder type 1;
- Current usage of psychotropic medication;
- Failure to respond to antidepressant medication in current episode;
- Electroconvulsive therapy for the treatment of the current episode of depression;
- Participants undergoing any form of face-to-face structured psychological treatment during the study;
- Clinically significant abnormal values for liver function tests, clinical chemistry, urine drug screen, blood pressure measurement and ECG. A participant with a clinical abnormality or parameters outside the reference range for the population being studied may be included only if the Investigator considers that the finding is unlikely to introduce additional risk factors and will not interfere with the study procedures;
- History of stimulant abuse (lifetime; e.g. amphetamine, cocaine), or of alcohol abuse within one year or of alcohol dependence within the lifetime;
- History of, or current medical conditions which in the opinion of the investigator may interfere with the safety of the participant or the scientific integrity of the study, including epilepsy/seizures, brain injury, severe hepatic or renal disease, severe gastro-intestinal problems, Central Nervous System (CNS) tumors, severe neurological problems (like Parkinson’s; blackouts requiring hospitalisation);
- Medical conditions that may alter the hemodynamic parameters underlying the BOLD signal (e.g., inadequately treated hypertension, diabetes mellitus), if attending Research Visit One;
- Clinically significant risk of suicide;
- Current pregnancy (as determined by urine pregnancy test taken during Screening and First Dose visits), breastfeeding or planning a pregnancy during the course of the study;
- Participant not willing to use a suitable method of contraception for 30 days after receiving study drug treatment;
- Any contraindication to MRI scanning (e.g. metal objects in body, pacemakers, significant claustrophobia, pregnancy), if attending Research Visit One;
- Participants with Body Mass Index (BMI) outside the 18 to 36 kg/m2 range at the Screening Visit.
- Night-shift working or recent travel involving significant change of timezones;
- Excessive caffeine consumption, i.e., consumption higher than 8 cups of standard caffeinated drinks (tea, instant coffee) or higher than 6 cups of stronger coffee or other drinks containing methylxanthines such as coca cola or Red Bull per day;
- Participation in a psychological or medical study involving the use of medication within the last 3 months;
- Previous participation in a study using the same, or similar, emotional processing tasks;
- Smoker > 10 cigarettes per day or similar levels of tobacco consumption in other forms.
- Participant received prescribed medication within 28 days prior to Visit 2 (apart from the contraceptive pill). Participants who have taken prescription medication may still be entered into the study, if, in the opinion of the Investigator, the medication received will not interfere with the study procedures or compromise safety;
- Participant received non-prescription medication, including supplements such as vitamins and herbal supplements within 48 hours prior to Visit 2 (apart from paracetamol). Participants who have taken non-prescription medication may still be entered into the study, if, in the opinion of the Investigator, the medication received will not interfere with the study procedures or compromise safety;
- Participant with a known hypersensitivity to PF-04995274, citalopram or any other serotonergic agents;
- Participant with planned medical treatment within the study period that might interfere with the study procedures;
- Participant who is unlikely to comply with the clinical study protocol or is unsuitable for any other reason, in the opinion of the Investigator.
- Participant with Covid-19 symptoms or a household member with Covid-19 symptoms

## Description of additional emotional cognition tasks

**Emotional Categorisation Task (ECAT):** Negative (e.g. “domineering”, “untidy”, “hostile”) or positive (e.g. “cheerful”, “honest”, “optimistic”) personality descriptions are presented on screen and participants are asked to indicate whether they would like or dislike to be described as each of these characteristics, with a button-press.

**Emotional Recall Task (EREC):** Participants are asked to write down as many of the words as they can remember from the ECAT. Incorrect words are latter classified by two independent researchers as positively or negatively valenced.

**Emotional Recognition Memory Task (EMEM):** Participants are presented with positive (e.g. “domineering”, “untidy”, “hostile”) and negative (e.g. “cheerful”, “honest”, “optimistic”) personality describing words. They are asked to indicate for each word whether it was presented in the ECAT, with a button press.

**Faces Dot Probe Task (FDOT):** Two faces are presented vertically on the computer screen and are promptly followed by a pair of dots. The participant is asked to indicate whether the dots are vertically or horizontally aligned, with a button press. Some of the faces will have an emotional expression (e.g. fearful or happy).

**Emotion potentiated startle (EPS):** an electromyography (EMG) is used to measure eye-blink responses to a burst of white noise while participants are presented with pictorial stimuli that are either positively valenced, negatively valenced or neutral.

**Probabilistic instrumental learning task (PILT), adapted from Pessiglione et al.** (**2006**): Participants are asked to win as much money as possible, by selecting symbols associated with the highest probability of winning money and lowest probability of losing money. On each trial, participants are presented with two pairs of symbols: one pair is associated with win outcomes (win £0.20 or no change) and the other with loss outcomes (lose £0.20 or no change). Each symbol in the pair has reciprocal probabilities (70% or 30%) of either outcome occurring and is randomly positioned either to the left or the right of a central fixation cross.

## Additional details about blinding

To achieve blinding between the three groups, each participant received 2 bottles, one containing 3 tablets and the other 1 capsule. Specifically, if they were assigned to the PF-04995274 treatment, they received 3 tablets of PF-04995274 (15mg in total) plus one capsule containing placebo. If they were assigned to the citalopram treatment, they received 1 encapsulated citalopram tablet (20 mg) plus 3 placebo tablets. Lastly, if participants were assigned to receive placebo, they received 3 placebo tablets plus 1 one capsule containing placebo.

## Data cleaning of emotion potentiated startle data

For emotion potentiated startle data, two researchers independently a) distinguished startle blink response from noise and decided whether a response could have been seen, had one occurred, excluding trials if a response could not be seen and b) determined if there was a blink response or if the trial should be recorded as a non-response. If there was disagreement, a third researcher made a final decision.

## Additional details about fMRI data acquisition

fMRI data was acquired with a multiband echo-planar imaging (EPI) sequence of 72 T-2 weighted slices covering the whole brain (echo time (TE) = 30 ms; repetition time (TR) = 1200 ms; flip angle 65°, field of view 216 mm, slice thickness 2 mm, multiband acceleration factor 4, PAT (GRAPPA) factor, voxel dimension 2mm isotropic, acquisition time 6min 28s), along with an acquired map of the field distortions to correct for intensity bias (echos at 4.92 and 7.38 ms, TR=590ms, flip angle = 46°). Additional high-resolution T1-weighted structural scans were acquired using a gradient echo sequence (TR 1900ms, TE 3.97ms, flip angle 8°, field of view 192mm, voxel dimension 1 mm isotropic, acquisition time 5min 31s) to allow later registration of the fMRI data into standard space.

## Additional details about fMRI pre-processing

Pre-processing involved various steps designed to reduce noise-related variability in the data and to improve the validity of the statistical analysis. Each participant’s functional imaging data underwent the following steps: (1) Removal of non-brain structures using BET, (2) motion correction using MCFLIRT, (3) spatial smoothing using a Gaussian kernel of FWHM 5 mm, (4) grand-mean intensity normalisation of the entire 4D dataset by a single multiplicative factor and high-pass temporal filtering cut-off = 90 s (Gaussian-weighted least-squares straight-line fitting, with sigma = 45 s), and (5) B0 unwarping using fieldmap rads and magnitude images for distortion correction.

## Supplementary Results

Self-reported affect – Table S1.

Self-reported affect – Table S1
| Visual Analogue Scale | PF-04995274, N = 31<sup>1</sup> | Citalopram, N = 30<sup>1</sup> | Placebo, N = 29<sup>1</sup> | F statistic | P value | Partial eta squared (η<sup>2</sup>) |
| --- | --- | --- | --- | --- | --- | --- |
| Current happiness | 46.97 (18.39) | 43.27 (18.03) | 45.31 (16.78) | Citalopram vs. placebo – F(1,57)= 0.202 | 0.66 | 0.004 |
|  |  |  |  | PF-04995274 vs. placebo – F(1,58) =0.133 | 0.72 | 0.002 |
| Current sadness | 29.72 (24.09) | 35.16 (24.61) | 34.14 (18.65) | Citalopram vs. placebo - F(1,57)= 0.033 | 0.86 | 0.0006 |
|  |  |  |  | PF-04995274 vs. placebo - F(1,58)= 0.623 | 0.43 | 0.01 |
| Current hostility | 9.34 (13.17) | 11.57 (16.84) | 11.55 (12.90) | Citalopram vs. placebo - F(1,57)= 0.00 | 0.99 | <0.0001 |
|  |  |  |  | PF-04995274 vs. placebo - F(1,58)= 0.431 | 0.51 | 0.007 |
| Current alertness | 35.30 (18.57) | 39.98 (19.31) | 44.16 (18.95) | Citalopram vs. placebo - F(1,57)=0.71 | 0.404 | 0.01 |
|  |  |  |  | PF-04995274 vs. placebo - F(1,58)=3.35 | 0.07 | 0.05 |
| Current anxiety | 29.05 (26.11) | 33.67 (25.97) | 38.60 (21.81) | Citalopram vs. placebo – F(1,57)=0.62 | 0.43 | 0.01 |
|  |  |  |  | PF-04995274 vs. placebo - F(1,58)=2.34 | 0.13 | 0.04 |
| Current calmness | 49.51 (18.32) | 51.23 (19.25) | 50.81 (19.45) | Citalopram vs. placebo – F(1,58)=0.007 | 0.93 | 0.0001 |
|  |  |  |  | PF-04995274 vs. placebo – F(1,58)=0.071 | 0.791 | 0.0012 |
| <sup>1</sup>Mean, averaged across final research visit measures (SD) |  |  |  |  |  |  |

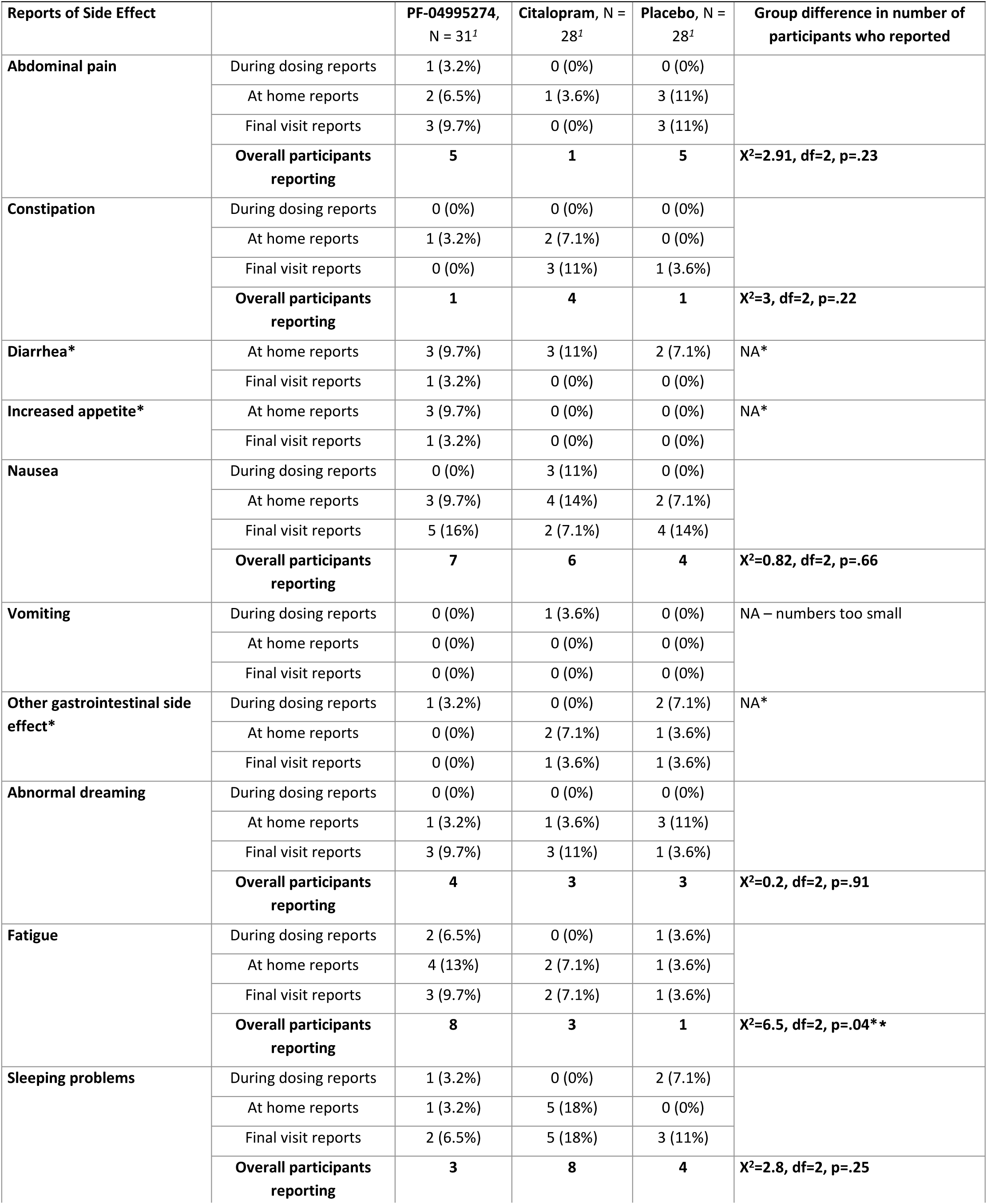
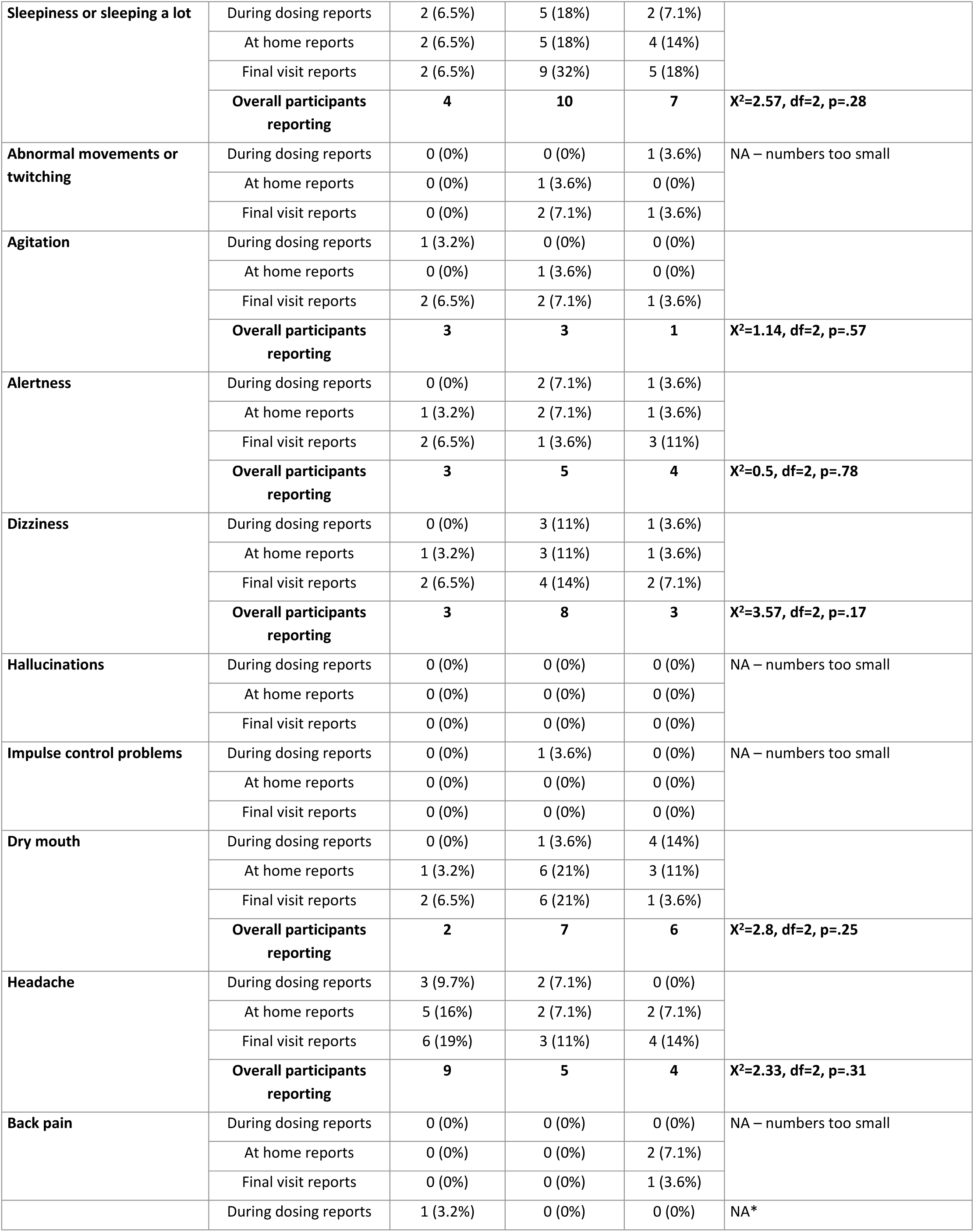
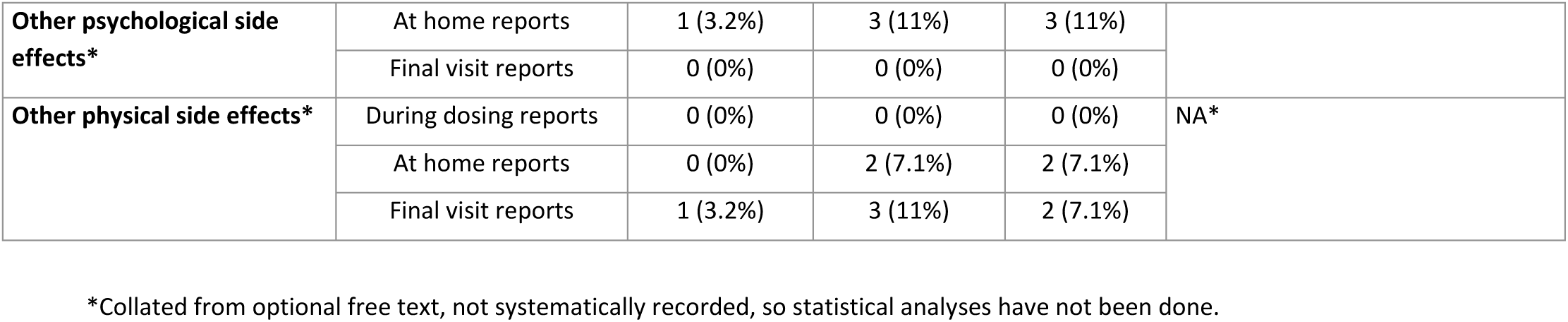
Side-effects – Table S2.

## ECAT, EREC, and EMEM

On the ECAT, EREC and EMEM, when comparing citalopram to placebo, there was no main effect of medication group (ECAT accuracy, F(1,112)= 1.308, p =0.255, ηp2=0.01; ECAT reaction time, F(1,112) = 0.782, p=0.378, ηp2=0.007; EREC correct words recalled, F(1,114)= 0.091, p =0.763, ηp2=0.0008; EREC incorrect words recalled, F(1,114)= 0.267, p =0.606, ηp2=0.002; EMEM total hits, F(1,110)=0.003, p=0.955, ηp2=0.00003; EMEM total correct rejections, F(1,110)=0.928, p=0.33747, ηp2=0.008; EMEM reaction time, F(1,110)=0.040, p=0.841, ηp2=0.0004).

There was also no interaction between valence and medication group (ECAT accuracy, F(1,112)= 1.574, p =0.212, ηp2=0.01; ECAT reaction time, F(1,112) = 0.150, p=0.699, ηp2=0.001; EREC correct words recalled, F(1,114)= 0.184, p=0.668, ηp2=0.002; EREC incorrect words recalled, F(1,114)= 0.002, p =0.967, ηp2=0.00001; EMEM total hits, F(1,110)=0.031, p=0.860, ηp2=0.0003; EMEM total correct rejections, F(1,110)=0.285, p=0.59483, ηp2=0.003; EMEM reaction time, F(1,110)=0.058, p=0.811, ηp2=0.0005).

Similarly, when comparing the PF-04995274 group to placebo, there was no main effect of medication group (ECAT accuracy, F(1,114)= 0.009, p =0.924, ηp2=0.00008; ECAT reaction time, F(1,114) = 0.002, p=0.966, ηp2=0.00002; EREC incorrect words recalled, F(1,116)= 0.259, p =0.612, ηp2=0.002; EMEM total hits, F(1,114)=1.062, p=0.305, ηp2=0.009; EMEM total correct rejections, F(1,114)=0.240, p=0.62517, ηp2=0.002; EMEM reaction time, F(1,114)=0.158, p=0.692, ηp2=0.001);

the only exception was a main effect of medication group on total correct words recalled in the EREC, with the PF-04995274 group recalling fewer words (F(1,116)= 5.321, p =0.0228, ηp2=0.04).

There was also no interaction between valence and medication group (ECAT accuracy, F(1,114)= 0.006, p =0.936, ηp2=0.00006; ECAT reaction time, F(1,114) = 0.221, p=0.639, ηp2=0.002; EREC correct words recalled, F(1,116)= 0.005, p =0.944, ηp2=0.00004; EREC incorrect words recalled, F(1,116)= 0.566, p =0.453, ηp2=0.005; EMEM total hits, F(1,114)=0.106, p=0.746, ηp2=0.0009; EMEM total correct rejections, F(1,114)=0.581, p=0.447, ηp2=0.005; EMEM reaction time, F(1,114)=0.072, p=0.790, ηp2=0.0006).

All results remained the same in sensitivity analyses, except that a main effect of medication group on EMEM total hits became significant, with the PF-04995274 group identifying less hits than the placebo group (F(1,108)=5.078, p=0.0263, ηp2=0.04).

## FDOT

There was no main effect of valence or mask. Neither the citalopram group nor the PF-04995274 group showed a significant main effect of medication (citalopram vs placebo, F(1,224)=0.052, p=0.820, ηp2=0.0002; PF-04995274 vs placebo, F(1,228)=0.949, p=0.331, ηp2=0.004), an interaction between medication group and valence (citalopram vs placebo, F(1,224)=0.149, p=0.700, ηp2=0.0007; PF-04995274 vs placebo, F(1,228)=0.864, p=0.354, ηp2=0.004), or an interaction between medication group, valence and mask (citalopram vs placebo, F(1,224)=1.809, p=0.180, ηp2=0.008; PF-04995274 vs placebo, F(1,228)=0.003, p=0.953, ηp2=0.00002). The results of these analyses did not change in sensitivity analyses.

## EPS

Across the whole sample, there was no evidence of the expected emotion potentiation – the main effect of image valence was non-significant (p>0.15) for amplitude (raw or Z scored VMax) and latency (TMax, p>0.75), and the largest amplitudes were seen for neutral stimuli. This pattern and lack of significance remained true within the placebo group as well.

When comparing citalopram to placebo on raw amplitude, there was a main effect of valence (F(2, 21)=3.76, p=0.040, ηp2=0.26), with mean amplitude for neutral being highest, followed by negative and then positive valence. There was also an interaction between group and valence (F(2,21)=4.08, p= 0.032, ηp2=0.28), with those on citalopram having greater amplitudes than placebo for neutral and negative images, and reduced amplitudes for positive images. In sensitivity analyses, the main effect of emotion became non-significant (F(2,20)= 0.99, p=0.39, ηp2=0.09), but the interaction remained (F(2,20)= 3.55, p=0.048, ηp2=0.26). When comparing PF-04995274 to placebo on raw amplitude, there was no significant interaction between group and valence (p>0.18).

When comparing citalopram to placebo on z-scored amplitude, there was no main effect of valence or interaction between group and valence (p>0.12). The same was true when comparing PF-04995274 to placebo (p>0.15).

When comparing citalopram to placebo on latency, there was a main effect of group (F(1,21)=12.36, p=0.002, ηp2=0.37), with citalopram showing shorter latency, but no interaction with valence (F(2,21)=0.202, p=0.82, ηp2=0.02). This remained true in sensitivity analyses. When comparing PF-04995274 to placebo on latency, there was no main effect of group or interaction between group and valence (p>0.2).

## PILT

Neither the citalopram group nor the PF-04995274 group showed a significant main effect of medication on choice of optimal symbol in win trials (citalopram vs placebo, F(1,53)=1.006, p=0.32, ηp2=0.02; PF-04995274 vs placebo, F(1,54)=0.095, p=0.759, ηp2=0.002), or in loss trials (citalopram vs placebo, F(1,53)=0.451, p=0.505, ηp2=0.008; PF-04995274 vs placebo, F(1,54)=0.01, p=0.922, ηp2=0.0002). The results of these analyses did not change in sensitivity analyses.

## fMRI behavioural results

Overall, the mean percentage accuracy for gender classification was 85.6% (sd=7.09, range 65.48-95.24%). For both groups, there was no main effect of group or interaction between group and valence, on percentage accuracy (citalopram vs placebo, main effect F(1,92)=1.126, p=0.219, ηp2=0.01, interaction F(1,92)=0.017, p=0.896, ηp2=0.0001; PF-04995274 vs placebo, main effect F(1,100)=0.821, p=0.367, ηp2=0.008, interaction F(1,100)=0.357, p=0.552, ηp2=0.003).

Overall, the mean reaction time was 642ms (sd=13, range 441-916ms). When comparing the citalopram and placebo groups, there was a significant main effect of medication group on reaction time (F(1,92)=12.914, p=0.0005, ηp2=0.12), with the citalopram group showing faster mean reaction times than placebo (m=603ms, sd=12.2 vs m=683ms, sd=9.8) but no interaction with valence of face (F(1,92)=0.821, p=0.367, ηp2=0.008). When comparing the PF-04995274 group to placebo, there was no significant main effect of medication (F(1,100)=2.981, p=0.087, ηp2=0.03), and no interaction between medication group and valence (F(1,100)=0.197, p=0.658 ηp2=0.02).

## fMRI data – task effects

When comparing mean activation to emotional faces (across fearful and happy faces) to activation during fixation in whole brain analysis, across all groups, there were significant clusters covering occipital pole, right cerebral cortex, post central gyrus, angular gyrus, precentral gyrus, cingulate gyrus, para-cingulate gyrus, left and right putamen, insular cortex, left thalamus, and left and right amygdala. ROI analyses found consistent activation in both amygdala, orbitofrontal cortex and medial-frontal cortex. These results are consistent with previous reports using the same task, suggesting that the task was successful in probing emotional processing. When comparing fear to happy i.e. looking at differential activation related to face valence, there were three significant clusters for fear > happy, two in cerebellum and one in the fusiform gyrus, and six significant clusters for fear < happy, including paracingulate gyrus, fusiform gyrus, middle and superior frontal gyrus, and brain stem. In ROI analyses, there was no significant differential activation of either amygdala, or the orbitofrontal cortex, but there was consistent significant activation associated with happy>fear in medial-frontal cortex.

fMRI whole brain analyses – Table S3.

fMRI whole brain analyses – Table S3
| Cluster number | Size (voxels) | F statistic (valence * contrast) | p-value | Z-max location (MNI) | Main regions involved in cluster | Mean fear – mean happy BOLD signal change % |
| --- | --- | --- | --- | --- | --- | --- |
| 6 | 541 | Citalopram vs. placebo –<br>F(1,90)= 4.101<br>PF-04995274 vs. placebo –<br>F(1,100) = 0.170 | 0.046<br>0.681 | 48, 50, 46, | Angular gyrus and lateral occipital | Placebo = -0.043<br>Citalopram = -0.143<br>PF-04995274 = 0.005 |
| 5 | 466 | Citalopram vs. placebo –<br>F(1,90)= 7.604<br>PF-04995274 vs. placebo –<br>F(1,100) = 0.482 | 0.007<br>0.489 | -40, -68, -48 | Cerebellum | Placebo = 0.118<br>Citalopram = -0.085<br>PF-04995274 = 0.069 |
| 4 | 393 | Citalopram vs. placebo –<br>F(1,90)= 7.211<br>PF-04995274 vs. placebo –<br>F(1,100) = 0.932 | 0.009<br>0.337 | -20, -86, -28 | Cerebellum, occipital fusiform, cerebral cortex | Placebo = 0.249<br>Citalopram = -0.206<br>PF-04995274 = 0.092 |
| 3 | 279 | Citalopram vs. placebo –<br>F(1,90)= 5.695<br>PF-04995274 vs. placebo –<br>F(1,100) = 0.597 | 0.019<br>0.441 | 68, -36, -8 | Middle temporal gyrus (posterior), superior temporal gyrus | Placebo = 0.058<br>Citalopram = -0.152<br>PF-04995274 = -0.016 |
| 2 | 191 | Citalopram vs. placebo –<br>F(1,90)= 4.408<br>PF-04995274 vs. placebo –<br>F(1,100) = 0.679 | 0.039<br>0.412 | 40, -70, -26 | Cerebellum, occipital fusiform, lateral occipital | Placebo = 0.138<br>Citalopram = -0.113<br>PF-04995274 = 0.052 |
| 1 | 116 | Citalopram vs. placebo –<br>F(1,90)= 4.71<br>PF-04995274 vs. placebo –<br>F(1,100) = 0.436 | 0.033<br>0.511 | 16, -82, -20 | Occipital fusiform, lingual gyrus | Placebo = 0.111<br>Citalopram = -0.105<br>PF-04995274 = 0.055 |

## Footnotes

1 HAM-6 includes items on depressed mood, guilt, work and activities, psychomotor retardation, psychic anxiety, and general somatic symptoms.

2 One FERT dataset for a participant in the placebo group was missing.

## References

1. Rush AJ, Trivedi MH, Wisniewski SR, Nierenberg AA, Stewart JW, Warden D, et al. Acute and longer-term outcomes in depressed outpatients requiring one or several treatment steps: a STAR*D report. Am J Psychiatry. 2006 Nov;163(11):1905–17.

2. Sharp T, Collins H. Mechanisms of SSRI Therapy and Discontinuation. Curr Top Behav Neurosci. 2024;66:21–47.

3. Balon R. SSRI-Associated Sexual Dysfunction. AJP. 2006 Sep;163(9):1504–9.

4. Colwell MJ, Tagomori H, Chapman S, Gillespie AL, Cowen PJ, Harmer CJ, et al. Pharmacological targeting of cognitive impairment in depression: recent developments and challenges in human clinical research. Transl Psychiatry. 2022 Nov 17;12(1):1–16.

5. GBD 2019 Diseases and Injuries Collaborators. Global burden of 369 diseases and injuries in 204 countries and territories, 1990-2019: a systematic analysis for the Global Burden of Disease Study 2019. Lancet. 2020 Oct 17;396(10258):1204–22.

6. Godlewska BR, Harmer CJ. Cognitive neuropsychological theory of antidepressant action: a modern-day approach to depression and its treatment. Psychopharmacology. 2021 May 1;238(5):1265–78.

7. Roiser JP, Elliott R, Sahakian BJ. Cognitive mechanisms of treatment in depression. Neuropsychopharmacology. 2012 Jan;37(1):117–36.

8. Harmer CJ, O’Sullivan U, Favaron E, Massey-Chase R, Ayres R, Reinecke A, et al. Effect of acute antidepressant administration on negative affective bias in depressed patients. Am J Psychiatry. 2009 Oct;166(10):1178–84.

9. Godlewska BR, Browning M, Norbury R, Cowen PJ, Harmer CJ. Early changes in emotional processing as a marker of clinical response to SSRI treatment in depression. Transl Psychiatry. 2016 Nov;6(11):e957–e957.

10. Ma Y. Neuropsychological mechanism underlying antidepressant effect: a systematic meta-analysis. Mol Psychiatry. 2015 Mar;20(3):311–9.

11. Williams LM, Korgaonkar MS, Song YC, Paton R, Eagles S, Goldstein-Piekarski A, et al. Amygdala Reactivity to Emotional Faces in the Prediction of General and Medication-Specific Responses to Antidepressant Treatment in the Randomized iSPOT-D Trial. Neuropsychopharmacology. 2015 Sep;40(10):2398–408.

12. Post A, Smart TS, Krikke-Workel J, Dawson GR, Harmer CJ, Browning M, et al. A Selective Nociceptin Receptor Antagonist to Treat Depression: Evidence from Preclinical and Clinical Studies. Neuropsychopharmacology. 2016 Jun;41(7):1803–12.

13. Murphy SE, Cates AN de, Gillespie AL, Godlewska BR, Scaife JC, Wright LC, et al. Translating the promise of 5HT4 receptor agonists for the treatment of depression. Psychological Medicine. 2021 May;51(7):1111–20.

14. Köhler-Forsberg K, Dam VH, Ozenne B, Sankar A, Beliveau V, Landman EB, et al. Serotonin 4 Receptor Brain Binding in Major Depressive Disorder and Association With Memory Dysfunction. JAMA Psychiatry. 2023 Apr 1;80(4):296–304.

15. Lucas G, Rymar VV, Du J, Mnie-Filali O, Bisgaard C, Manta S, et al. Serotonin4 (5-HT4) Receptor Agonists Are Putative Antidepressants with a Rapid Onset of Action. Neuron. 2007 Sep 6;55(5):712–25.

16. Mendez-David I, David DJ, Darcet F, Wu MV, Kerdine-Römer S, Gardier AM, et al. Rapid Anxiolytic Effects of a 5-HT4 Receptor Agonist Are Mediated by a Neurogenesis-Independent Mechanism. Neuropsychopharmacology. 2014 May;39(6):1366–78.

17. Pascual-Brazo J, Castro E, Díaz Á, Valdizán EM, Pilar-Cuéllar F, Vidal R, et al. Modulation of neuroplasticity pathways and antidepressant-like behavioural responses following the short-term (3 and 7 days) administration of the 5-HT4 receptor agonist RS67333. Int J Neuropsychopharmacol. 2012 Jun 1;15(5):631–43.

18. Harmer CJ, Shelley NC, Cowen PJ, Goodwin GM. Increased Positive Versus Negative Affective Perception and Memory in Healthy Volunteers Following Selective Serotonin and Norepinephrine Reuptake Inhibition. AJP. 2004 Jul;161(7):1256–63.

19. Hamilton M. A RATING SCALE FOR DEPRESSION. J Neurol Neurosurg Psychiatry. 1960 Feb;23(1):56–62.

20. Beck AT, Steer RA, Ball R, Ranieri W. Comparison of Beck Depression Inventories –IA and –II in psychiatric outpatients. J Pers Assess. 1996 Dec;67(3):588–97.

21. Snaith RP, Hamilton M, Morley S, Humayan A, Hargreaves D, Trigwell P. A scale for the assessment of hedonic tone the Snaith-Hamilton Pleasure Scale. Br J Psychiatry. 1995 Jul;167(1):99–103.

22. Spielberger CD. State-Trait Anxiety Inventory for Adults [Internet]. 2012 [cited 2025 Mar 7]. Available from: https://doi.apa.org/doi/10.1037/t06496-000

23. Eysenck SBG, Eysenck HJ, Barrett P. A revised version of the psychoticism scale. Personality and Individual Differences. 1985 Jan 1;6(1):21–9.

24. Watson D, Clark LA, Tellegen A. Development and validation of brief measures of positive and negative affect: the PANAS scales. J Pers Soc Psychol. 1988 Jun;54(6):1063–70.

25. Bond A, Lader M. The use of analogue scales in rating subjective feelings. British Journal of Medical Psychology. 1974;47(3):211–8.

26. Thomas JM, Higgs S, Dourish CT. Test–retest reliability and effects of repeated testing and satiety on performance of an Emotional Test Battery. Journal of Clinical and Experimental Neuropsychology. 2016 Apr 20;38(4):416–33.

27. de Cates AN, Gillespie AL, Scaife J, Martens MAG, Carson J, et al. Effects on hippocampal activity following 5-HT4 receptor agonism in unmedicated patients with depression: the RESTAND study. medRxiv, 333049.

28. de Cates AN, Martens MAG, Wright LC, Gould van Praag CD, Capitão LP, Gibson D, et al. The Effect of the 5-HT4 Agonist, Prucalopride, on a Functional Magnetic Resonance Imaging Faces Task in the Healthy Human Brain. Front Psychiatry. 2022;13:859123.

29. Tassone VK, Gholamali Nezhad F, Demchenko I, Rueda A, Bhat V. Amygdala biomarkers of treatment response in major depressive disorder: An fMRI systematic review of SSRI antidepressants. Psychiatry Res Neuroimaging. 2024 Mar;338:111777.

30. Pizzagalli DA, Roberts AC. Prefrontal cortex and depression. Neuropsychopharmacology. 2022 Jan;47(1):225–46.

31. Zimmerman M, Martinez JH, Young D, Chelminski I, Dalrymple K. Severity classification on the Hamilton depression rating scale. Journal of Affective Disorders. 2013 Sep 5;150(2):384–8.

32. Klein-Flügge MC, Bongioanni A, Rushworth MFS. Medial and orbital frontal cortex in decision-making and flexible behavior. Neuron. 2022 Sep 7;110(17):2743–2770.

33. Nord CL, Barrett LF, Lindquist KA, Ma Y, Marwood L, Satpute AB, et al. Neural effects of antidepressant medication and psychological treatments: a quantitative synthesis across three meta-analyses. The British Journal of Psychiatry. 2021 Oct;219(4):546–550.

34. Murphy SE, Wright LC, Browning M, Cowen PJ, Harmer CJ. A role for 5-HT4 receptors in human learning and memory. Psychological Medicine. 2020 Dec;50(16):2722–30.

35. Masdrakis VG, Markianos M, Baldwin DS. Apathy associated with antidepressant drugs: a systematic review. Acta Neuropsychiatrica. 2023 Aug;35(4):189–204.

36. Cates AN de, Harmer CJ, Harrison PJ, Cowen PJ, Emmanuel A, Travis S, et al. Association between a selective 5-HT4 receptor agonist and incidence of major depressive disorder: emulated target trial. The British Journal of Psychiatry. 2024 Sep;225(3):371–8.

37. de Cates AN, Wright LC, Martens MAG, Gibson D, Türkmen C, Filippini N, et al. Déjà-vu? Neural and behavioural effects of the 5-HT4 receptor agonist, prucalopride, in a hippocampal-dependent memory task. Transl Psychiatry. 2021 Oct 4;11(1):1–9.

